# Prioritizing the Role of Major Lipoproteins and Subfractions as Risk Factors for Peripheral Artery Disease

**DOI:** 10.1101/2021.01.11.21249148

**Authors:** Michael G. Levin, Verena Zuber, Venexia M. Walker, Derek Klarin, Julie Lynch, Rainer Malik, Aaron W. Aday, Leonardo Bottolo, Aruna D. Pradhan, Martin Dichgans, Kyong-Mi Chang, Daniel J. Rader, Philip S. Tsao, Benjamin F. Voight, Dipender Gill, Stephen Burgess, Scott M. Damrauer, on behalf of the VA Million Veteran Program

**Affiliations:** Division of Cardiovascular Medicine, University of Pennsylvania Perelman School of Medicine, Philadelphia, PA; Department of Medicine, University of Pennsylvania Perelman School of Medicine, Philadelphia, PA; Corporal Michael J. Crescenz VA Medical Center, Philadelphia, PA; MRC Biostatistics Unit, School of Clinical Medicine, University of Cambridge, Cambridge, UK; Department of Epidemiology and Biostatistics, Imperial College London, London, UK; Medical Research Council Integrative Epidemiology Unit, University of Bristol, Bristol, UK; Department of Surgery, University of Pennsylvania Perelman School of Medicine, Philadelphia, PA; Malcolm Randall VA Medical Center, Gainesville, FL; Department of Surgery, University of Florida, Gainesville, FL; VA Informatics and Computing Infrastructure, Department of Veterans Affairs, Salt Lake City Health Care System, Salt Lake City, UT, USA; University of Utah School of Medicine, Salt Lake City, UT, USA; Institute for Stroke and Dementia Research, University Hospital of Ludwig-Maximilians-University, Munich, Germany; Vanderbilt Translational and Clinical Cardiovascular Research Center, Division of Cardiovascular Medicine, Vanderbilt University Medical Center, Nashville, TN; Department of Medical Genetics, School of Clinical Medicine, University of Cambridge, Cambridge, UK; The Alan Turing Institute, London, UK; Division of Preventive Medicine, Brigham and Women’s Hospital, Harvard Medical School, Boston, MA; Division of Cardiovascular Medicine, VA Boston Medical Center, Boston, MA; Institute for Translational Medicine and Therapeutics, University of Pennsylvania Perelman School of Medicine, Philadelphia, PA; Palo Alto VA Healthcare System, Palo Alto, CA; Department of Medicine, Division of Cardiovascular Medicine, and Stanford Cardiovascular Institute, Stanford University, Palo Alto, CA; Department of Genetics, University of Pennsylvania Perelman School of Medicine, Philadelphia, PA; Department of Systems Pharmacology and Translational Therapeutics, University of Pennsylvania Perelman School of Medicine, Philadelphia, PA; Department of Epidemiology and Biostatistics, Imperial College London, UK; Clinical Pharmacology and Therapeutics Section, Institute for Infection and Immunity, St. George’s, University of London, London, UK; Novo Nordisk Research Centre Oxford, Old Road Campus, Oxford, UK; BHF Cardiovascular Epidemiology Unit, School of Clinical Medicine, University of Cambridge, Cambridge, UK

**Keywords:** Peripheral Artery Disease, Lipoproteins, Genetics, Mendelian Randomization

## Abstract

**Background:** Circulating lipid and lipoprotein levels have consistently been identified as risk factors for atherosclerotic cardiovascular disease (ASCVD), largely on the basis of studies focused on coronary artery disease (CAD). The relative contributions of specific lipoproteins to risk of peripheral artery disease (PAD) have not been well-defined. Here, we leveraged large scale genetic association data to identify genetic proxies for circulating lipoprotein-related traits, and employed Mendelian randomization analyses to investigate their effects on PAD risk.

**Methods:** Genome-wide association study summary statistics for PAD (Veterans Affairs Million Veteran Program, 31,307 cases) and CAD (CARDIoGRAMplusC4D, 60,801 cases) were used in the Mendelian Randomization Bayesian model averaging (MR-BMA) framework to prioritize the most likely causal major lipoprotein and subfraction risk factors for PAD and CAD. Mendelian randomization was used to estimate the effect of apolipoprotein B lowering on PAD risk using gene regions that proxy potential lipid-lowering drug targets. Transcriptome-wide association studies were performed to identify genes relevant to circulating levels of prioritized lipoprotein subfractions.

**Results:** ApoB was identified as the most likely causal lipoprotein-related risk factor for both PAD (marginal inclusion probability 0.86, p = 0.003) and CAD (marginal inclusion probability 0.92, p = 0.005). Genetic proxies for ApoB-lowering medications were associated with reduced risk of both PAD (OR 0.87 per 1 standard deviation decrease in ApoB, 95% CI 0.84 to 0.91, p = 9 × 10^−10^) and CAD (OR 0.66, 95% CI 0.63 to 0.69, p = 4 × 10^−73^), with a stronger predicted effect of ApoB-lowering on CAD (ratio of ORs 1.33, 95% CI 1.25 to 1.42, p = 9 × 10^−19^). Among ApoB-containing subfractions, extra-small VLDL particle concentration (XS.VLDL.P) was identified as the most likely subfraction associated with PAD risk (marginal inclusion probability 0.91, p = 2.3 × 10^−4^), while large LDL particle concentration (L.LDL.P) was the most likely subfraction associated with CAD risk (marginal inclusion probability 0.95, p = 0.011). Genes associated with XS.VLDL.P and L.LDL.P included canonical ApoB-pathway components, although gene-specific effects varied across the lipoprotein subfractions.

**Conclusion:** ApoB was prioritized as the major lipoprotein fraction causally responsible for both PAD and CAD risk. However, diverse effects of ApoB-lowering drug targets and ApoB-containing lipoprotein subfractions on ASCVD, and distinct subfraction-associated genes suggest possible biologic differences in the role of lipoproteins in the pathogenesis of PAD and CAD.

## INTRODUCTION

Atherosclerotic cardiovascular disease (ASCVD) is the most common cause of morbidity and mortality worldwide (1). Most research has focused on ASCVD in the coronary arteries (coronary artery disease, CAD). However, peripheral artery disease (PAD) represents another common and often underrecognized manifestation of ASCVD that is also associated with significant morbidity and mortality, affecting more than 5% of the global adult population (2,3). Dyslipidemia has been a long-established risk factor for ASCVD, with the strongest evidence derived from large studies primarily focused on CAD endpoints. Although low-density lipoprotein cholesterol (LDL-C) reducing medications like statins are commonly employed in the prevention and treatment of PAD, evidence on the relationship between LDL-C and PAD risk has been inconsistent (4). Observational studies with modest event rates have suggested that components of the atherogenic dyslipidemia profile (elevated levels of triglyceride-rich lipoproteins, small LDL-C, and the ratio of total cholesterol to HDL-cholesterol [HDL-C] along with low concentrations of HDL-C) may be more strongly associated with PAD than CAD, although the relative contribution of the major circulating lipoproteins (LDL-C, HDL-C, triglycerides, apolipoprotein B [ApoB], and apolipoprotein A1 [ApoA1]) and associated lipoprotein subfractions to PAD specifically has remained poorly defined (4–7).

Over the past 15 years, genome-wide association studies have identified hundreds of genetic loci associated with ASCVD traits, major lipoproteins, and related subfractions (8–12). The large datasets arising from these studies include genetic associations estimated in hundreds of thousands of participants. An array of analytic methods enable the analysis of these genetic datasets to provide insights into the underlying biology of diseases. Mendelian randomization (MR) uses genetic variants as instrumental variables to infer the effect of an exposure on an outcome, under the assumption that genetic associations with the outcome are mediated via the exposure for selected variants (13). MR has been used to implicate ApoB as an important risk factor for CAD, and to validate the effects of drug targets on disease outcomes (14–19). Integrating genetic data with gene transcription datasets in transcriptome-wide association studies (TWAS) has been used to identify tissue-level gene expression associations with disease (such as between hepatic expression of *SORT1* and risk of CAD) (20).

We aimed to integrate large-scale genetic datasets to 1) prioritize the role of circulating lipoproteins and subfractions on PAD risk; 2) identify genes that may represent novel lipoprotein-pathway targets in the prevention and treatment of PAD; and 3) estimate the effects of current/potential lipid-lowering medications on PAD risk.

## METHODS

### Study Population and Outcomes

Our primary outcome was PAD. Genetic associations with PAD were derived from a 2019 genome-wide association study by Klarin et al (10). Full summary data are available by application to dbGaP (phs001672.v4.p1). This study included 31,307 PAD cases (24,009 European-ancestry, 5,373 African-ancestry, 1,925 Hispanic-ancestry) and 211,753 controls among participants of the Veterans Affairs Million Veteran Program (21), which recruited individuals aged ≥19 from Veterans Affairs Medical Centers across the United States. PAD diagnoses were ascertained from electronic health records using International Classification of Diseases (ICD)-9/10 and Current Procedural Terminology (CPT) codes. Genetic associations were performed separately by ancestry groups using logistic regression adjusted for age, sex, and five ancestry-specific genetic principal components, and then combined using an inverse-variance weighted fixed-effects method.

CAD was included as an outcome in our analysis to help contextualize the PAD results, as most of the observational, Mendelian randomization, and randomized control trial data relating to ASCVD have focused on CAD outcomes. Genetic associations with CAD were derived from the CARDIoGRAMplusC4D 1000 Genomes GWAS (11). This is a meta-analysis of 48 studies, including 60,801 CAD cases and 123,504 controls primarily of European ancestries (77%), including a combination incident and prevalent CAD among the cases.

### Prioritizing the Role of Major Lipoprotein-related Traits and Lipoprotein Subfractions on PAD

We performed a variable selection method in a multivariable MR framework to prioritize the causal lipoprotein determinants of the outcomes. Multivariable MR extends the basic MR framework to include multiple exposures in one joint model (22,23), which is particularly relevant when considering highly correlated traits like blood lipoprotein-related traits as exposures. In order to rank and select the likely causal lipoprotein characteristics for PAD, we employed an extension of multivariable MR called Mendelian randomization Bayesian model averaging (MR-BMA) (14), a Bayesian approach for prioritizing causal exposures in a two-sample multivariable MR setting. MR-BMA performs variable selection by evaluating models with all possible combinations of lipoprotein-related traits as exposures and computing the posterior probability for each model that the model contains the true causal risk factors. The evidential support for each exposure (the marginal inclusion probability) is derived from the sum of all posterior probabilities of the models where the specific exposure was included. We removed influential variants based on the Cook’s distance and outliers based on the *q*-statistic as previously recommended (14). An empirical permutation procedure was used to calculate *p-*values which are adjusted for multiple testing using the Benjamini-Hochberg false discovery rate (FDR) procedure.

We performed two MR-BMA analyses as illustrated in **Figure 1A**. First, to identify causal relationships between major lipoprotein-related traits and PAD, our exposures of interest were circulating lipoproteins: low-density lipoprotein cholesterol (LDL-C), high-density lipoprotein cholesterol (HDL-C); their primary constituent apolipoproteins: ApoA1 and ApoB; and triglycerides (TG) (**Figure 1A**). Second, in order to investigate whether the predicted effect of ApoB-lowering on PAD may be influenced by specific lipoprotein subfractions, we performed a further MR-BMA analysis focusing on ten ApoB-containing lipoprotein subfractions (**Figure 1A**). For comparison, we also performed these analyses for CAD.

**Figure 1:**
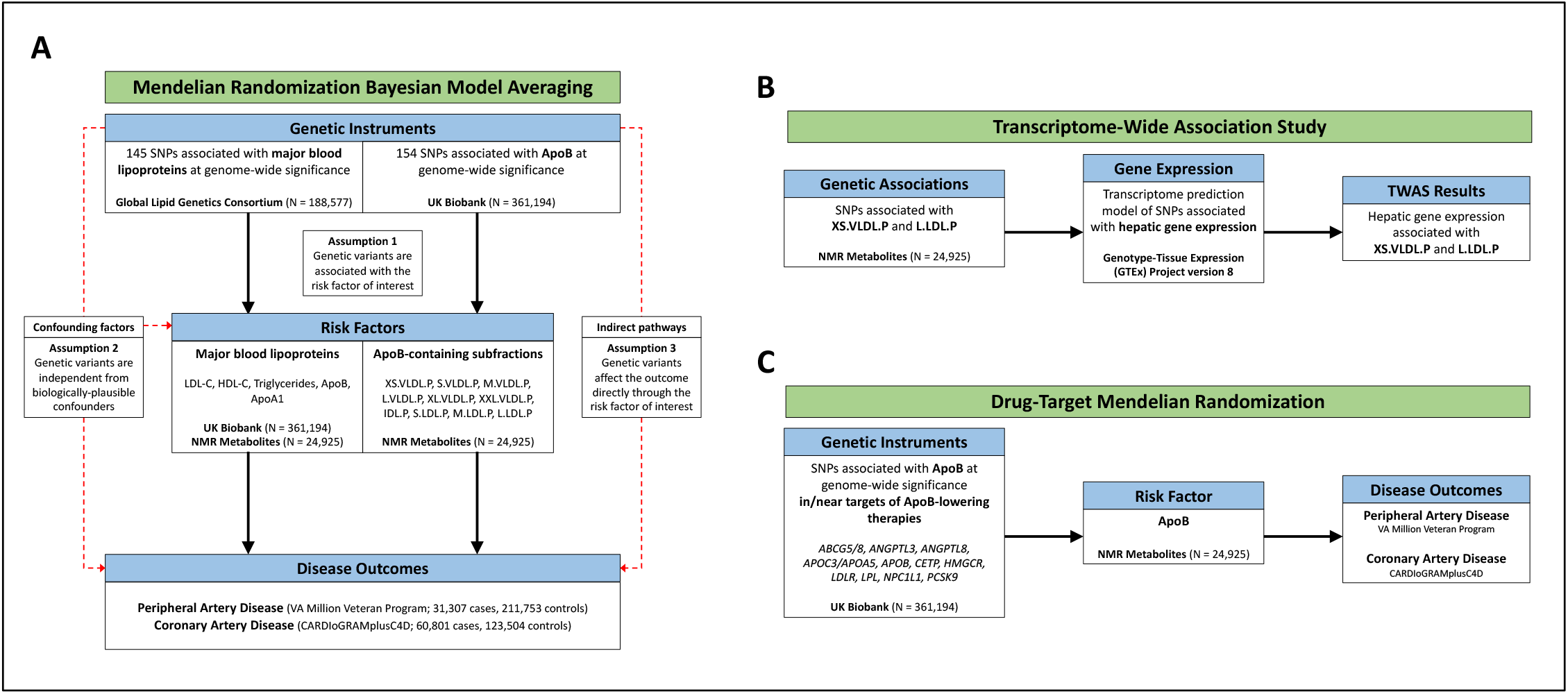
Overview of Risk Factor Prioritization, Drug-Target, and TWAS Analyses. Overview of main analyses. A) Risk factor prioritization was performed using MR-BMA to prioritize the contribution of 1) major lipoproteins, and 2) ApoB-containing subfractions to PAD and CAD risk. The primary MR assumptions are denoted, with red dashed lines representing violations of the MR assumptions. B) A Transcriptome-Wide Association Study integrating gene expression and genetic association data was performed to identify putative genes involved in regulation of the prioritized ApoB-containing subfractions. C) Drug-target MR was performed to identify the effect of genes encoding targets of ApoB-lowering medications on PAD and CAD outcomes.

The instrumental variables consisted of 145 independent (r^2^ < 0.001 in the 1000 Genomes European-ancestry Reference Panel) genetic variants associated with any major lipoprotein-related trait (total cholesterol, LDL-cholesterol, HDL-cholesterol, or triglycerides) at a genome-wide significance level (p < 5 × 10^−8^) in the Global Lipids Genetics Consortium, a GWAS comprising 60 individual studies of primarily European-ancestry participants (8). Genetic associations with circulating levels of major lipoprotein-related traits in blood were estimated in the UK Biobank study based on 361,194 European-ancestry participants (http://www.nealelab.is/uk-biobank/). Genetic associations were adjusted for age, sex, and twenty principal components. Genetic associations with lipoprotein subfractions were estimated from a GWAS of circulating lipoproteins and subfractions in 24,925 European-descent participants (9). In this dataset, lipoproteins and subfractions were measured using nuclear magnetic resonance (NMR) spectroscopy. Genetic associations were adjusted for age, sex, and the first 10 genetic principal components. As a replication analysis, we repeated the analysis for major lipoprotein-related traits using genetic associations from the NMR dataset.

### Transcriptome-Wide Association Study

After prioritizing the role of lipoprotein subfractions in PAD, we next sought to identify genes associated with those subfractions, which may ultimately serve as therapeutic targets (**Figure 1B**). To identify genes associated with circulating levels of lipoprotein subfractions, transcriptome-wide association studies (TWAS) were performed using S-PrediXcan (20). This tool enables the integration of tissue-level expression quantitative trait loci (eQTL) datasets with GWAS summary statistics to prioritize genes associated with traits of interest. As the liver plays a critical role in lipoprotein metabolism, we obtained a pre-trained transcriptome prediction model for liver gene expression derived from the Genotype-Tissue Expression (GTEx) Project version 8 (http://predictdb.org). Predicted liver gene expression and GWAS summary statistics for lipoprotein subfractions were then correlated using S-PrediXcan to identify genes significantly associated with circulating levels of lipoprotein subfractions. The significance of differences in the effect of each gene on each outcome were determined by: 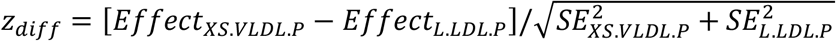 with p-values derived from the normal distribution. We performed gene ontology enrichment analysis using ShinyGO to identify Gene Ontology Biological Processes significantly associated with the genes prioritized by S-PrediXcan (24). Finally, we performed a combined multivariate and collapsing test to investigate the effect of rare damaging mutations in genes prioritized by the TWAS on risk of PAD among participants of the UK Biobank who underwent whole exome sequencing (25) (**Supplemental Methods**).

### Genetically-proxied ApoB-lowering and PAD Risk

To predict the impact of ApoB-lowering on PAD and CAD risk, we performed further MR analyses (**Figure 1C**). We performed gene-based analyses using variants associated with ApoB in gene regions that proxy specific lipid-lowering drugs (licensed or proposed), and polygenic analyses using all such variants. Genetic variants associated with ApoB levels at genome-wide significance (p < 5 × 10^−8^) were identified from the from the UK Biobank (http://www.nealelab.is/uk-biobank/) and pruned at r^2^<0.1 to exclude highly correlated variants. This set of variants was further narrowed into two biologically-informed sets. First, we examined variants located in or near (+/-200kb) genes encoding previously-identified regulators of ApoB metabolism (*ABCG5/8, ANGPTL3, ANGPTL4, ANGPTL8, APOC3/APOA5, APOB, CETP, DGAT, HMGCR, LDLR, LPL, MTTP, NPC1L1, PCSK9*, and *PPARA*), representing the targets of current or proposed therapeutics (16). Next, we examined variants in or near (+/-200kb) genes associated with extra-small VLDL particle concentration (XS.VLDL.P) in the TWAS analysis (FDR q < 0.05). Genetic associations with ApoB were taken from the NMR dataset (9) to avoid winner’s curse and sample overlap. MR estimates were obtained from the random effects inverse-variance weighted method performed using the *MendelianRandomisation* package in R, accounting for linkage disequilibrium correlation among variants using the 1000 Genomes Phase 3 European reference panel.

### Statistical Analysis

For the main MR-BMA analyses of major lipoprotein-related traits and PAD, FDR correction was performed to account for multiple testing, with FDR-corrected q < 0.05 set as the predetermined significance threshold. For the MR-BMA analysis of lipoprotein subfractions, the Nyholt procedure of effective tests was used to account for the strong correlation among the subfractions, with a multiple testing adjusted p-value of 0.05 set as the significance threshold (26). For the drug target MR analysis, p < 0.05 was the predetermined significance threshold. For the TWAS and gene ontology enrichment analyses, the false-discovery rate was used to account for multiple testing, with FDR-corrected p < 0.05 set as the predetermined significance threshold. All statistical analyses were performed using R version 4.0.3 (R Foundation for Statistical Computing (https://www.R-project.org/). This study is reported in accordance with the STROBE guidelines for reporting observational studies (27).

## RESULTS

### Prioritizing the Role of Major Lipoprotein-related Traits and Lipoprotein Subfractions on PAD

In the MR-BMA analysis for major lipoprotein-related traits, ApoB was the top-ranked risk factor for PAD (marginal inclusion probability 0.86, p = 0.003). (**Table 1A**; Supplemental Tables 4-6). There was also nominal evidence for ApoA1 (marginal inclusion probability 0.53, p = 0.025) and HDL-C (marginal inclusion probability 0.47, p = 0.031); however, after accounting for multiple testing, these exposures were not prioritized for inclusion in the model. In the replication analysis, where genetic association estimates for the five major lipoprotein-related traits were derived from the NMR metabolite GWAS, ApoB was again the top-ranked risk factor for PAD with a marginal inclusion probability of 0.68 (p = 0.001) (**Supplemental Tables 7-9**). There was also nominal evidence for HDL-C (marginal inclusion probability 0.501, p = 0.039), which again no longer persisted after correcting for multiple testing. Similarly, ApoB was identified as the prioritized risk factor for CAD in the primary (marginal inclusion probability 0.92, p = 0.005) (**Table 1B**) and replication (marginal inclusion probability 0.80, p = 0.004) analyses, in keeping with the previously established role of ApoB in CAD (**Supplemental Tables 10-11**) (15,16). These results provide strong, consistent support for the role of ApoB as the primary lipoprotein risk factor for PAD and CAD.

**Table 1:** Prioritization of Causal Risk Factors Amongst Major Lipoproteins. Ranking of most likely causal exposures amongst major lipoproteins for peripheral artery disease and coronary artery disease prioritized using multivariable Mendelian randomization in the Mendelian randomization Bayesian Model Averaging (MR-BMA) framework. False discovery rate (FDR) corrected *p*-values are derived in an empirical permutation procedure. LDL-C: low-density lipoprotein cholesterol, HDL-C: high-density lipoprotein cholesterol, ApoA1: apolipoprotein A1, ApoB: apolipoprotein B.

| **A) Peripheral Artery Disease** |  |  |  |
| --- | --- | --- | --- |
| **Exposure** | **Marginal inclusion probability** | **Uncorrected *p*-value** | **FDR corrected *p*-value** |
| ApoB | 0.856 | 0.003 | 0.015 |
| ApoA1 | 0.529 | 0.025 | 0.052 |
| HDL-C | 0.467 | 0.031 | 0.052 |
| LDL-C | 0.461 | 0.168 | 0.210 |
| Triglycerides | 0.238 | 0.386 | 0.386 |
| **B) Coronary Artery Disease** |  |  |  |
| **Exposure** | **Marginal inclusion probability** | **Uncorrected *p*-value** | **FDR corrected *p*-value** |
| ApoB | 0.922 | 0.005 | 0.027 |
| HDL-C | 0.457 | 0.022 | 0.054 |
| LDL-C | 0.33 | 0.746 | 0.746 |
| ApoA1 | 0.284 | 0.123 | 0.206 |
| Triglycerides | 0.152 | 0.580 | 0.726 |

In the MR-BMA analysis for lipoprotein subfractions, extra-small VLDL particle concentration (XS.VLDL.P) was prioritized as the primary ApoB-containing risk factor for PAD (marginal inclusion probability 0.91, p = 2.3 × 10^−4^) (**Table 2A; Supplemental Table 15**). In contrast, large LDL particle concentration (L.LDL.P) was prioritized as the primary ApoB-containing risk factor for CAD (marginal inclusion probability 0.95, p = 0.011) (**Table 2B; Supplemental Table 16**).

**Table 2:** Prioritization of Causal Risk Factors Amongst Apolipoprotein B-containing Lipid Subfractions. Ranking of most likely causal exposures amongst apolipoprotein B-containing lipid subfractions for peripheral artery disease and coronary artery disease prioritized using multivariable Mendelian randomization in the Mendelian randomization Bayesian Model Averaging (MR-BMA) framework. False discovery rate (FDR) corrected *p*-values are derived using the Nyholt procedure of effective tests to account for the strong correlation among the subfractions. XXL.VLDL.P: Extra-extra large very-large density lipoprotein particles, XL.VLDL.P: Extra large very-large density lipoprotein particles, L.VLDL.P: Large very-large density lipoprotein particles, M.VLDL.P: Medium very-large density lipoprotein particles, S.VLDL.P: Small very-large density lipoprotein particles, XS.VLDL.P: Extra small very-large density lipoprotein particles, L.LDL.P: Large large-density lipoprotein particles, M.LDL.P: Medium large-density lipoprotein particles, S.LDL.P: Small large-density lipoprotein particles, IDL.P: Intermediate-density lipoprotein particles.

| **A) Peripheral Artery Disease** |  |  |  |
| --- | --- | --- | --- |
| **Exposure** | **Marginal inclusion probability** | **Uncorrected *p*-value** | **FDR corrected *p*-value** |
| XS.VLDL.P | 0.912 | 0.0023 | 0.00115 |
| IDL.P | 0.078 | 0.963 | 1.000 |
| L.LDL.P | 0.023 | 0.999 | 1.000 |
| S.VLDL.P | 0.022 | 0.901 | 1.000 |
| M.LDL.P | 0.021 | 0.999 | 1.000 |
| XL.VLDL.P | 0.021 | 0.996 | 1.000 |
| S.LDL.P | 0.020 | 1.000 | 1.000 |
| XXL.VLDL.P | 0.017 | 1.000 | 1.000 |
| L.VLDL.P | 0.014 | 0.999 | 1.000 |
| M.VLDL.P | 0.014 | 0.997 | 1.000 |
| **B) Coronary Artery Disease** |  |  |  |
| **Exposure** | **Marginal inclusion probability** | **Uncorrected *p*-value** | **FDR corrected *p*-value** |
| L.LDL.P | 0.613 | 0.008 | 0.040 |
| M.LDL.P | 0.382 | 0.554 | 1.000 |
| IDL.P | 0.071 | 0.614 | 1.000 |
| S.LDL.P | 0.045 | 0.981 | 1.000 |
| XS.VLDL.P | 0.036 | 0.294 | 1.000 |
| S.VLDL.P | 0.027 | 0.765 | 1.000 |
| M.VLDL.P | 0.023 | 0.900 | 1.000 |
| L.VLDL.P | 0.016 | 0.948 | 1.000 |
| XL.VLDL.P | 0.015 | 0.986 | 1.000 |
| XXL.VLDL.P | 0.014 | 0.950 | 1.000 |

### Identification of Genes Associated with ApoB-containing Lipoprotein Subfractions

Having prioritized XS.VLDL.P and L.LDL.P as important ApoB-containing lipoprotein subfractions for PAD and CAD respectively, we explored whether specific genes may influence the circulating levels of these subfractions. Given the key role of the liver in lipoprotein metabolism, we integrated hepatic gene expression data from the Genotype-Tissue Expression (GTEx) Project with the XS.VLDL.P and L.LDL.P GWAS summary statistics to identify genes associated with circulating levels of each ApoB-containing lipoprotein subfraction.

Using TWAS, we identified 31 genes associated with XS.VLDL.P and 23 genes associated with L.LDL.P, for a total of 40 unique genes (FDR <0.05 for either subfraction) (**Supplemental Tables 17-18**). Of these, We identified 17 genes uniquely associated with XS.VLDL.P levels, 9 genes uniquely associated with L.LDL.P levels, and 14 genes associated with circulating levels of both subfractions (**Figure 2A-B**; Supplemental Tables 17-18). There was overall strong correlation between the estimated effects of gene expression on circulating XS.VLDL.P and L.LDL.P levels (Pearson correlation = 0.80, p < 2.2 × 10^−16^). As expected, genes associated with these lipoprotein subfractions were significantly enriched for membership in cholesterol metabolism and related pathways (**Supplemental Table 19**). We also identified unique genes associated with each subfraction (FDR < 0.05 for either subfraction). Among the genes associated with both subfractions were several canonical genes involved in lipoprotein metabolism, including *PCSK9, ABCG8, LIPC*, and *APOA5*. The 17 genes uniquely associated with XS.LDL.P levels were enriched for clusters of biological processes involving triglyceride-rich lipoprotein metabolism (**Supplemental Table 20**), while the 9 genes uniquely associated with L.LDL.P levels were not significantly enriched for specific biological processes. Of the 40 genes associated with either lipoprotein subfraction, we identified potentially heterogenous effects on circulating lipoprotein subfraction levels. For example, *NLRC5, LIPC, CETP, APOA5, USP1, ANGPTL3*, and *ATGC4* expression was predicted to more strongly impact circulating XS.VLDL.P levels in comparison to L.LDL.P (**Figure 2C**).

**Figure 2:**
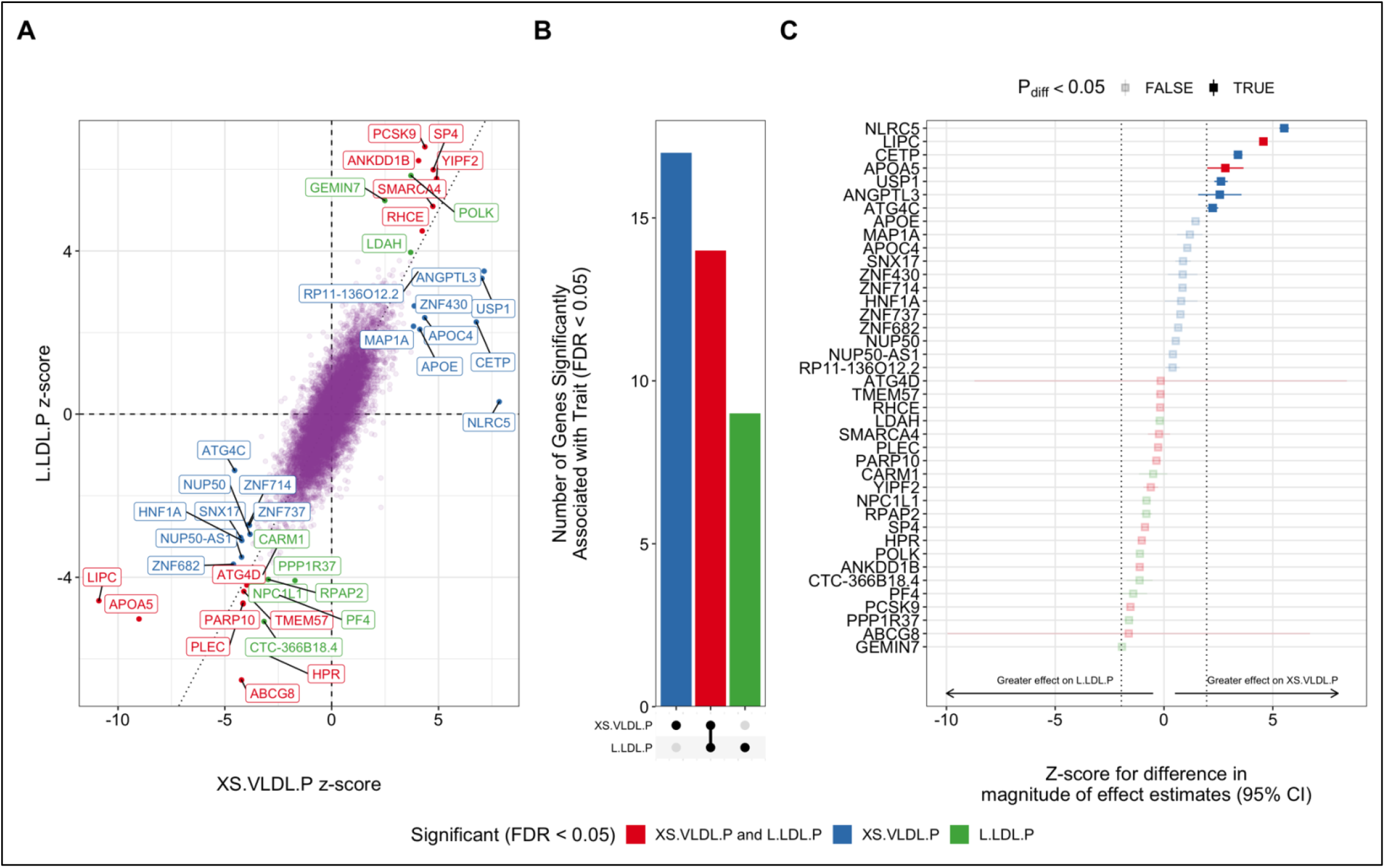
Genes Associated with Circulating Levels of XS. VLDL.P and L.LDL.P Lipoprotein Subfractions. Transcriptome-wide association studies were performed integrating liver gene-expression data from GTEX v8 with GWAS summary statistics for XS.VLDL.P and L.LDL.P to identify genes associated with circulating levels of each lipoprotein subfraction. **A)** Genes significantly (FDR < 0.05) associated with either subfraction are labeled, and colors represent the subfraction associations. **B)** The bar plot depicts the number of unique and shared genes between the two subfractions. **C)** The forest plot depicts the z-score for the difference in the effect magnitude for each gene on each subfraction. Dotted lines represent z-scores of +/-1.96, with point estimates outside this range representing significant (p_diff_ < 0.05) differential effects. Errors represent 95% confidence intervals for the z-score.

To investigate the impact of these genes on PAD risk, we examined whether damaging mutations in XS.VLDL.P-associated genes might influence PAD risk among UK Biobank participants. We identified rare loss-of-function variants in 29 of 31 XS.VLDL.P-associated genes among 154,584 UK Biobank participants (1,668 PAD cases and 152,916 controls). After accounting for multiple testing, only damaging variants in *SP4* were associated with prevalent PAD (FDR q = 0.049) (**Supplemental Table 21**).

### Genetically-predicted ApoB-lowering and PAD Risk

Finally, because Mendelian randomization has previously been utilized to predict the impact of current and proposed ApoB-lowering therapies on CAD risk (16), we sought to explore the effect of these treatments on PAD risk. We first performed polygenic and gene-based MR analyses to determine whether ApoB-associated genetic variants located within/near genes encoding these therapeutic targets were associated with risk of PAD. In polygenic analyses, genetically-proxied ApoB-lowering was associated with reduced risk of PAD (OR 0.87 per 1 standard deviation reduction in ApoB, 95% CI 0.84 to 0.91, p = 9 × 10^−9^) (**Figure 3A**). As a comparison, the association of genetically-proxied ApoB-lowering with CAD risk using the same genetic variants was greater (OR 0.66 per 1 standard deviation decrease in circulating ApoB, 95% CI 0.63 to 0.69, p = 4 × 10^−73^; ratio of ORs 1.33, 95% CI 1.25 to 1.42, p = 9 × 10^−19^).

**Figure 3:**
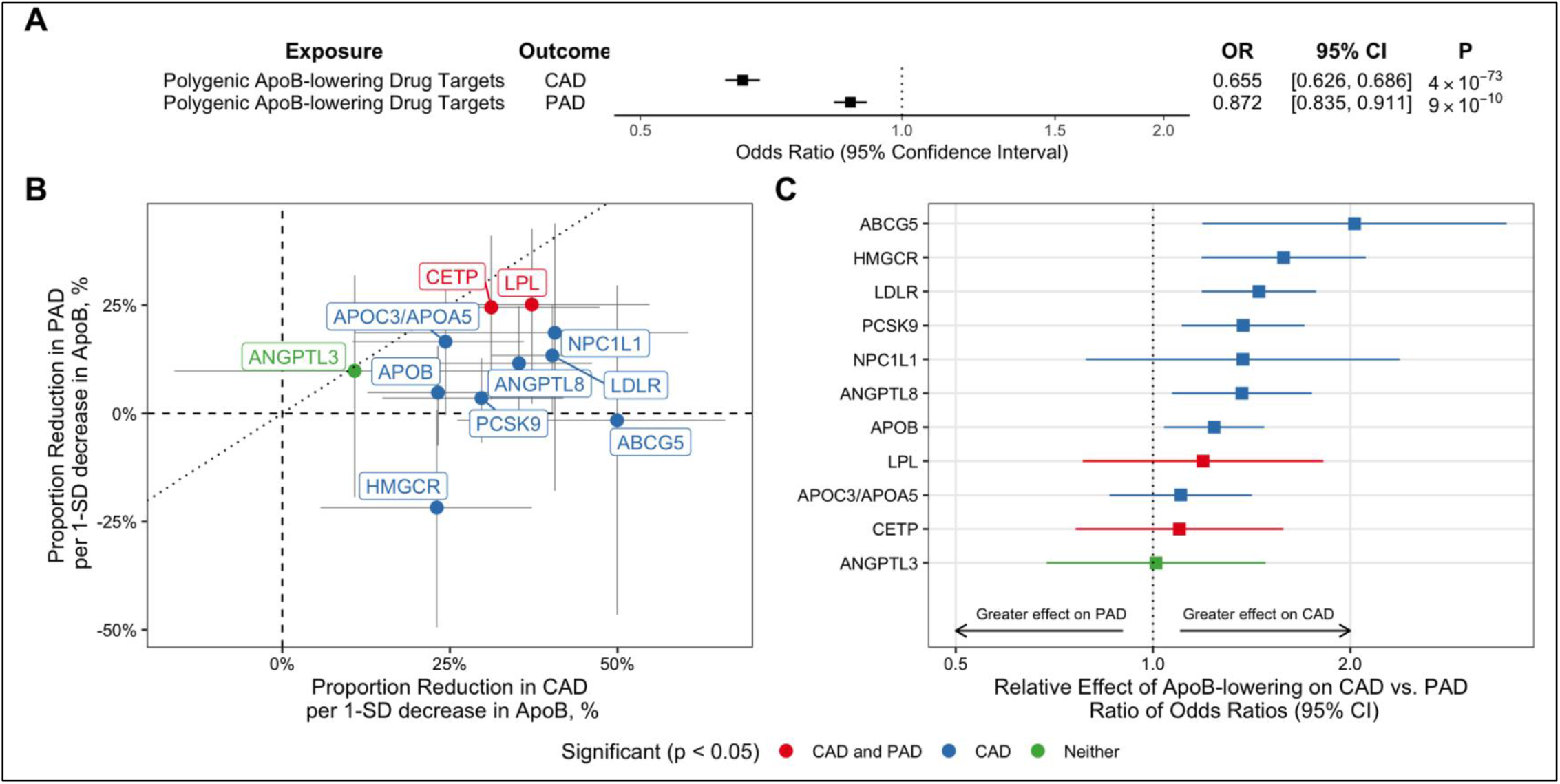
Associations Between Genetically-Predicted ApoB levels and PAD. Estimates represent associations between genetically-proxied ApoB and PAD or CAD risk, scaled to the change in disease risk per 1 standard deviation (1-SD) decrease in ApoB. Genetic variants used to proxy reductions in ApoB only included those located near/within genes important in ApoB metabolism. **A)** Polygenic analysis including all such variants. OR = odds ratio, CI = confidence interval. **B)** Gene-based analyses considering variants for each gene region. The dotted line with slope 1 represents the scenario where the association of genetically-proxied ApoB with disease risk is equal for both CAD and PAD. **C)** Relative effects of each gene region on CAD vs. PAD as determined by the ratio of odds ratios. Error bars represent 95% confidence intervals.

Next, we compared the associations between genetically-proxied ApoB-lowering and ASCVD outcomes in gene-based analyses, identifying potential heterogenous effects on PAD and CAD risk. We identified protective effects on CAD and/or PAD for 11 of 12 ApoB target genes (**Figure 3B**; **Supplemental Table 14**). While several associations did not achieve statistical significance for PAD specifically, associations were generally in the risk decreasing direction, with the exception of *HMGCR* locus which trended toward increased PAD risk (although 95% confidence intervals did not exclude a small protective effect on PAD). Consistent with the overall polygenic analysis, genetically-proxied ApoB-lowering at the *ABCG5, HMGCR, LDLR, PCSK9, ANGPTL8*, and *APOB* loci had significantly greater protective effects on CAD in comparison to PAD (ratio of ORs > 1, FDR < 0.05) (**Figure 3C**).

Finally, we performed polygenic and gene-specific MR analyses to explore whether XS.VLDL.P-associated genes identified in the TWAS analysis were associated with PAD risk. In the polygenic analysis, ApoB-lowering proxied by genetic variants located within/near XS.VLDL.P-associated genes was associated with reduced risk of PAD (OR 0.89 per 1 SD reduction in ApoB, 95% CI 0.86 to 0.92, p = 3 × 10^−11^) (**Figure 4**). In gene-specific analyses, ApoB-lowering at the *CETP, NLRC5*, and *YIPF2* loci were significantly associated with decreased PAD risk, while ApoB-lowering at the *ANKDD1B* locus was associated with increased PAD risk (**Figure 4**).

**Figure 4:**
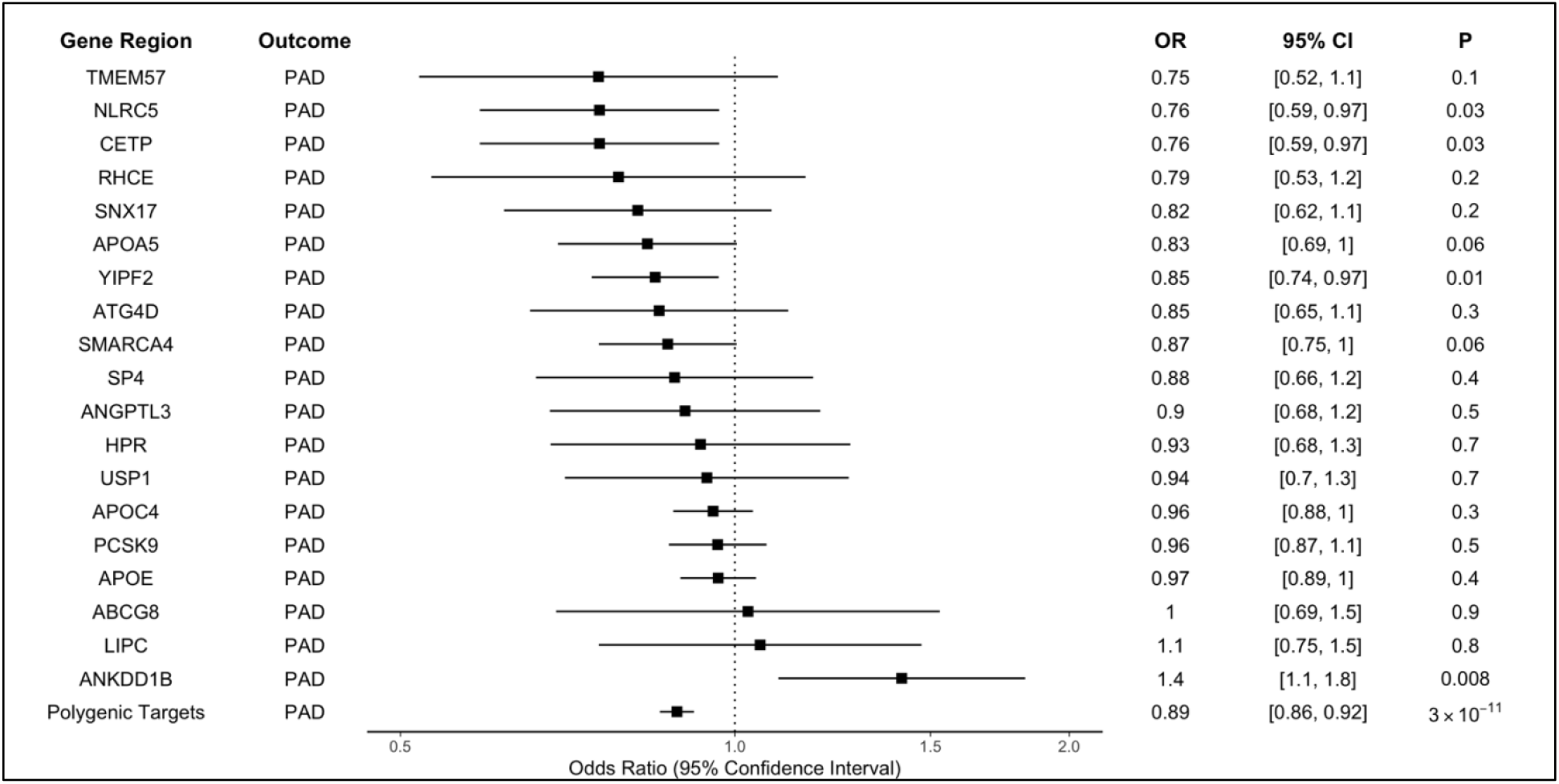
Associations between XS. VLDL.P-associated Genes and PAD. Polygenic and gene-specific MR were performed to estimate the association between the ApoB-lowering effect of XS.VLDL.P-associated genes and PAD. Gene-specific MR examining the impact of ApoB-associated variants in/near (+/-200kb) each XS.VLDL.P-associated gene, with Polygenic Targets denoting the aggregate impact of variants in/near these genes.

## DISCUSSION

We integrated several large genetic datasets and an array of statistical genetics, molecular epidemiology, and bioinformatic tools to uncover novel causal relationships between circulating lipoprotein-related traits and PAD, and compared these findings with CAD. First, we identified ApoB as the primary major circulating lipoprotein-related trait responsible for risk of PAD, similar to CAD. Next, we prioritized XS.VLDL.P as the ApoB-associated subfraction most strongly associated with PAD risk, in contrast to CAD where L.LDL.P was the most strongly associated lipoprotein subfraction. We identified genes involved in the regulation of important ApoB-containing lipoprotein subfractions, which may represent directed targets for novel PAD prevention and treatment strategies. Finally, we explored the impact of ApoB-lowering on PAD and uncovered the potential for the XS.VLDL.P pathway to be targeted to reduce PAD risk.

These results highlight similarities and differences in the roles of circulating lipoproteins for PAD and CAD. Our primary analysis identified ApoB as the major lipoprotein-related trait responsible for both PAD and CAD risk. This finding is consistent with a recent meta-analysis of 22 studies (including 1,892 PAD cases and 30,937 controls) which found significantly higher ApoB levels among PAD cases compared to controls (28). A nested case-control study within the Physicians’ Health Study (PHS) similarly identified baseline ApoB levels (in addition to several other lipid fractions) as a significant predictor of incident PAD (29). In contrast, a large observational study including 31,657 participants of 5 prospective Finnish cohorts did not detect an association between ApoB levels and incident PAD, but may have been limited by a low incidence of PAD (498 cases) and by defining PAD based on hospitalization codes (30). Similarly, while the Women’s Health Study (WHS) did not identify an association between baseline ApoB and incident PAD (5), differences in PAD case definitions, ascertainment, and demographics (incident PAD in WHS vs. prevalent in MVP; women in WHS vs. predominately men in MVP and PHS), may account for these differences. Our prioritization of ApoB as the most important lipoprotein-related risk factor for CAD is consistent with recent MR studies establishing ApoB as the primary risk factor for CAD (15,16). In the setting of strong epidemiologic and genetic correlation between CAD and PAD, it is not surprising that these two manifestations of ASCVD share ApoB as a common risk factor. However, we identified a stronger effect of ApoB on CAD than PAD, which has implications for risk stratification and treatment.

ApoB-containing particles exist on a spectrum of varying sizes, densities, and particle compositions (31), and identification of specific subfractions that contribute to different forms of atherosclerotic cardiovascular disease may have implications for pathophysiology, molecular mechanisms, risk stratification, and treatment. Variability in the distribution of ApoB within lipoprotein subfractions may contribute to differential risk of PAD compared to CAD. Indeed, clinical observations have suggested that Type III hyperlipoproteinemia (familial dysbetalipoproteinemia), a disorder specifically associated with VLDL remnant particles, may be a greater risk factor for PAD than for CAD (31). In our genetic analyses of ApoB-containing lipoprotein subfractions using MR-BMA, extra-small VLDL particle concentration (XS.VLDL.P) was the primary ApoB-containing subfraction contributing to PAD risk. In contrast, large LDL particle concentration (L.LDL.P) was the primary ApoB-containing subfraction contributing to CAD risk. These results are consistent with a recent observational study exploring the effect of circulating lipoproteins and metabolites on incident PAD and CAD among 31,657 participants of five prospective Finnish cohorts, which found a strong association between XS.VLDL.P and incident PAD, with no significant association between L.LDLP and incident PAD (30). The primary effect of XS.VLDL.P on PAD implicates an important role for triglyceride-rich lipoproteins and remnant particles in the pathogenesis of PAD. This stands in contrast to the effect of L.LDL.P on CAD, which suggests that LDL-associated lipoprotein fractions may play a more important role in the pathogenesis of coronary artery disease. The mechanisms by which different ApoB-containing subfractions contribute to ASCVD risk requires further study. Intriguingly, a prior MR analysis proposed a mechanism by which remnant particles causally increase inflammation (as measured by C-reactive protein level), whereas no inflammatory effect was detected in the setting of genetically-proxied elevations in LDL cholesterol (32).

While both PAD and CAD are manifestations of ASCVD, there are pathophysiologic differences between the two diseases that may provide a basis for targeted treatment strategies (33). Although ApoB represents a common lipoprotein risk factor for PAD and CAD, we identified differences in predicted response to ApoB-lowering treatment, underscoring potential differences in the role of circulating lipoproteins on these ASCVD outcomes. We demonstrate that while ApoB-lowering is expected to have favorable effects on both PAD and CAD risk, the relative benefit is expected to be significantly greater for CAD risk-reduction in comparison to PAD risk-reduction. Although large randomized controlled trials, genetic studies, and observational evidence have highlighted the importance of LDL-cholesterol and ApoB-lowering in reducing ASCVD outcomes overall, these studies have primarily focused on major adverse cardiovascular events and CAD outcomes (7,34,35). Our results reveal that the ApoB-lowering effect of several clinically approved and clinical trial-stage drug targets is predicted to differ between PAD and CAD, a finding which may have implications for both drug discovery and treatment paradigms. Although we identified varied effects across ApoB-lowering targets, these results should not at this point be used to guide treatment decisions, and MR and clinical trial estimates of treatment effects may vary (36). While overall these results support guideline recommendations for the use of ApoB-lowering medications to reduce PAD risk, our results also argue for PAD-specific outcomes to be measured in cardiovascular outcomes trials, as the absence of a treatment effect for CAD (or a combined endpoint) may not exclude PAD-specific effects (1,35). For example, The PROMINENT (Pemafibrate to Reduce Cardiovascular Outcomes by Reducing Triglycerides in Patients with Diabetes) Study will evaluate the impact of triglyceride-rich lipoprotein lowering on major cardiovascular events with adjudicated PAD events a secondary endpoint of the trial (37).

Finally, our polygenic and gene-based drug target analyses highlighted potential targets for directed treatment strategies. Although our TWAS analysis identified associations between several canonical genes involved in lipoprotein metabolism and circulating levels of both XS.VLDL.P and L.LDL.P, we also identified distinct genes associated with each lipoprotein subfraction which may have treatment implications. As lipid-lowering medications induce specific changes in the circulating lipoprotein profile (38), our results suggest that specific drug-target identification may play an important role in identifying PAD-focused treatments. For example, genes associated with circulating XS.VLDL.P were enriched in pathways related to triglyceride-rich lipoprotein metabolism. Although our polygenic and gene-specific MR analyses suggested that both currently available and proposed ApoB-lowering therapies would be expected to reduce PAD risk, we used MR to further highlight the XS.VLDL.P pathway as a potentially novel therapeutic target. Whether these genes and pathways represent pharmacologic targets that ultimately impact PAD outcomes warrants further study.

### Limitations

This study should be interpreted within the context of its limitations. First, this study focused on prevalent PAD outcomes ascertained from electronic health records. The effect of lipoprotein-related traits may vary across specific incident PAD outcomes including intermittent claudication, rest pain, tissue-loss, and amputation. Second, PAD outcomes were studied among primarily male participants of the Veterans Affairs Million Veteran Program, and although participants were of diverse ancestries, further studies among other populations are warranted to improve generalizability of these findings. Third, MR effect estimates reflect lifelong genetic exposures, and may not accurately reflect the magnitude of benefit of shorter-term pharmacologic interventions (39). Thus, our drug-target MR findings should not be used to guide clinical decisions regarding lipid-lowering therapies at this stage. Fourth, when correlated exposures exist within a common pathway, the MR-BMA method identifies the most proximate risk factor to the outcome. Circulating levels of each lipid fraction are composed of several subcomponents (for example, ApoB, triglycerides, and cholesterol are all found in VLDL, IDL, and LDL compartments and subfractions). Prioritization of ApoB over LDL-C for PAD and CAD risk does not nullify LDL-C as a causal risk factor, but indicates that effects of lipid-lowering therapies are likely to be proportional to the change in ApoB rather than LDL-C. Detailed MR analysis of other lipid subfractions may further prioritize additional PAD risk factors and identify additional therapeutic targets.

Overall, this analysis of large genetic datasets identified ApoB as the primary causal lipoprotein-related risk factor for PAD. Diverse effects of ApoB-lowering drug targets and ApoB-containing lipoprotein subfractions on PAD in comparison to CAD suggests possible biologic differences in the pathogenesis of these diseases, with gene expression analyses revealing potential targets for novel PAD therapies.

## Supporting information

MVP Collaborators

## Data Availability

The analytic code from this study is available from the corresponding author upon request. GWAS summary statistics for PAD are available by application in dbGaP (phs001672.v4.p1). GWAS summary statistics for GLGC lipids, UKB lipids, NMR lipids, and CARDIoGRAMplusC4D CAD are available for download from the IEU Open GWAS Project (https://gwas.mrcieu.ac.uk/).

## ACKNOWLEDGEMENTS

This research is based on data from the MVP, Office of Research and Development, Veterans Health Administration and was supported by award no. MVP003/MVP028 (I01-BX003362). A list of MVP collaborators is included in the supplement. This work was supported by US Department of Veterans Affairs grants IK2-CX001780 (SMD). This publication does not represent the views of the Department of Veterans Affairs or the United States government. DG is funded by the British Heart Foundation Centre of Research Excellence (RE/18/4/34215) at Imperial College London. SB is supported by a Sir Henry Dale Fellowship jointly funded by the Wellcome Trust and the Royal Society (Grant Number 204623/Z/16/Z).

## ETHICAL APPROVAL

This study was approved by the VA Central Institutional Review Board.

