## Supplemental Methods for "Prioritizing the Role of Major Lipoproteins and Subfractions as Risk Factors for Peripheral Artery Disease"

### SUPPLEMENTAL MATERIAL

#### SUPPLEMENTAL METHODS

##### UK BioBank Loss-of-Function Burden Analysis

To estimate the effect of damaging mutations in XS.VLDL.P-associated genes on risk of PAD, we performed a burden test among UK Biobank participants who underwent whole exome sequencing (WES). PAD was defined using ICD10 codes from death records and hospital episode stays (HES), ICD9 codes and OPCS4 codes, as previously described [1]. All individuals with >1 code were assigned a case status, whereas all other individuals were assigned a control status. In the complete UK Biobank dataset (N=502,336), we identified 6,329 unique PAD cases (1.26% cases in population). In the UK Biobank WES dataset (N=200,644), we identified 2,147 unique PAD cases (1.07% cases in population). We excluded from this dataset: i) individuals from non-British White ancestry and ii) individuals with excess heterozygosity. For burden analysis, we also excluded related individuals (up to 2$^{nd}$ degree; KING cutoff 0.0884) using PRIMUS [2] while retaining cases preferentially to controls (high_btrait option in PRIMUS). Our final dataset consisted of 1,668 cases and 152,916 controls (1.08% cases in population). For the 31 selected genes, we selected rare variants (MAF < 0.01) that are either predicted to be damaging (REVEL [3] score > 0.5) or predicted to exert a high-confidence loss-of-function effect using the LoFTEE plugin [4] in VEP [5]. We pooled those variants in a combined burden analysis using the CMC unidirectional burden test [6] implemented in the rvtests software [7]. The CMC test was performed using age at baseline, sex and 10 PCs as covariates.
